# Development and validation of risk prediction model for recurrent cardiovascular events among Chinese: P-CARDIAC model

**DOI:** 10.1101/2023.10.17.23297127

**Authors:** Yekai Zhou, Jiaxi Lin, Qiuyan Yu, Joseph Edgar Blais, Eric Yuk Fai Wan, Marco Lee, Emmanuel Wong, David Chung-Wah Siu, Vincent Wong, Esther Wai Yin Chan, Tak-Wah Lam, William Chui, Ian Chi Kei Wong, Ruibang Luo, Celine SL Chui

**Author notes:** co-first authors (Yekai Zhou, Jiaxi Lin). co-correspondence (Ruibang Luo, Celine SL Chui). Dr Ruibang Luo, Rm 301 Chow Yei Ching Building, The University of Hong Kong, Pokfulam Road, Pokfulam, Hong Kong Special Administration Region, China. Dr Celine Sze Ling Chui, 5/F Academic Building, 3 Sassoon Road, Pokfulam, Hong Kong Special Administration Region, China.

## Abstract

This study aimed to develop and validate a cardiovascular diseases (CVD) risk prediction model, Personalized CARdiovascular DIsease risk Assessment for Chinese (P-CARDIAC), for recurrent cardiovascular events using Machine-Learning technique.

Three cohorts of Chinese patients with established CVD in Hong Kong were included; Hong Kong Island cohort as the derivation cohort, whilst the Kowloon and New Territories cohorts were validation cohorts. The 10-year CVD outcome was a composite of diagnostic or procedure codes for coronary heart disease, ischaemic or haemorrhagic stroke, peripheral artery disease, and revascularization. We estimated incidence of recurrent CVD events for each cohort with reference to the total person-years of each cohort. Multivariate imputation with chained equations (MICE) and XGBoost were applied for the model development. The comparison with TRS-2°P and SMART2 used the validation cohorts with 1000 bootstrap replicates.

A total 48,799, 119,672 and 140,533 patients were included in the derivation and validation cohorts, respectively. A list of 125 risk variables were used to make predictions on CVD risk, of which, eight classes of medications were considered interactive drug use. Model performance in the derivation cohort showed satisfying discrimination and calibration with a C-statistic of 0·69. Internal validation showed good discrimination and calibration performance with C-statistic over 0·6. P-CARDIAC also showed better performance than TRS-2°P and SMART2.

Compared to other risk scores, P-CARDIAC enables to identify unique patterns of Chinese patients with established CVD. We anticipate that P-CARDIAC can be applied in various settings to prevent recurrent CVD events, thus reducing the related healthcare burden.

**Condensed Abstract:** A CVD risk prediction model named Personalized CARdiovascular DIsease risk Assessment for Chinese (P-CARDIAC), for recurrent cardiovascular events among Chinese adults using Machine-Learning technique was newly developed. It predicted 10-year CVD outcome including a composite of diagnostic or procedure codes for coronary heart disease, ischaemic or haemorrhagic stroke, peripheral artery disease, and revascularization by incidence of recurrent CVD. Model showed satisfying discrimination and calibration with a C-statistic of 0·69. P-CARDIAC also showed better performance than existing risk scores, such as TRS-2°P and SMART2. P-CARDIAC could help predict recurrent CVD risk and reduce the healthcare burden.

## Introduction

Cardiovascular diseases (CVD), including coronary heart disease and stroke, are the leading cause of non-communicable deaths globally, with an estimated 18·6 million fatalities recorded in 2019.^1,2^ CVD is also the leading cause of death and disease burden in China, contributing to 3·72 million deaths and total hospitalization costs of approximately US $10·7 billion in 2013.^3-5^ In Hong Kong, heart disease and cerebrovascular diseases are the third and fourth leading cause of deaths in 2021.^6^ However, according to a World Health Organization report, 80% of premature heart attacks and strokes are preventable.^7^

Some research groups advocate the use of risk prediction models on patients to identify those at high risk of CVD who are more likely to benefit from preventive strategies.^8-11^ The development and applicability of CVD risk prediction models are highly dependent on the ethnic and socioeconomic factors of the population of interest.^12^ Currently, there are several risk scores for recurrent CVD risk prediction among individuals with established CVD, including The Thrombolysis in Myocardial Infarction (TIMI) Risk Score for Secondary Prevention (TRS-2°P) and Secondary Manifestations of ARTerial disease (SMART2) risk score.^13,14^ These risk scores provide an estimated risk of recurrent CVD, and thus help provide early intervention to patients with less resource implications.^15^ However, these models are tailored to western populations, whose applicability to other ethnicities is uncertain. There has been limited validation of the influence of ethnicity on the application of CVD risk scores and these results are poorly calibrated for Asian populations in Southeast Asia.^16^ In addition, although treatment options such as lipid-modifying therapies are effective in secondary prevention among those with established CVD, the estimation of treatment effect is often not considered in current risk scores.^17-19^ Therefore, a risk prediction model specifically tailored to the Chinese population for secondary prevention, incorporating dynamic medication treatment with drugs proven to reduce CVD risk is of paramount importance to identify the means to reduce the CVD healthcare burden.

In this study, we developed and validated the Personalized CARdiovascular DIsease risk Assessment for Chinese (P-CARDIAC) among the Chinese population in Hong Kong using Machine-Learning (ML) technique. The ML technique has been used to identify patterns in large data sets to enable delivery of healthcare services by facilitating effective patient-provider decision-making.^20^ P-CARDIAC was developed to provide early intervention for patients at high risk of recurrent CVD by leveraging the rich data source of electronic health records (EHR). It estimates the 10 years of recurrent CVD risk for high-risk individuals with consideration of an array of risk variables captured in the EHR. We also validated the performance of P-CARDIAC, TRS-2°P and SMART2 on the representative study cohorts from Hong Kong, a city in Southeast Asia where over 90% of inhabitants are of Chinese ethnicity.^21^

## Methods

### Study cohorts

Three cohorts of patients with established CVD were identified based on geographical location of residence in Hong Kong (Hong Kong West Cluster, Hong Kong Island; Kowloon; New Territories). The Hong Kong Island (Hong Kong West Cluster) cohort was used for model derivation whilst the Kowloon and New Territories cohorts were used for model validation.

Patients were included if they had used any of the public healthcare services provided by the Hong Kong Hospital Authority (HA) since 2004 (inclusion and exclusion criteria detailed in Figure 1 and Supplementary Information 1). HA is a statutory body and the largest public healthcare provider of Hong Kong. It provides government subsidised primary, secondary and tertiary care to all residents, capturing over 70% of all hospitalisations in Hong Kong.^22^ Previous studies demonstrated high validity of the data source with a positive predictive value of 85% for myocardial infarction (MI) and 91% for stroke.^23^ The database was also used for over two hundred studies published in peer-reviewed journals, including cardiovascular diseases and cardiovascular drugs studies, ensuring the creditability of the data source for research purposes.^23-26^

**Figure 1.**
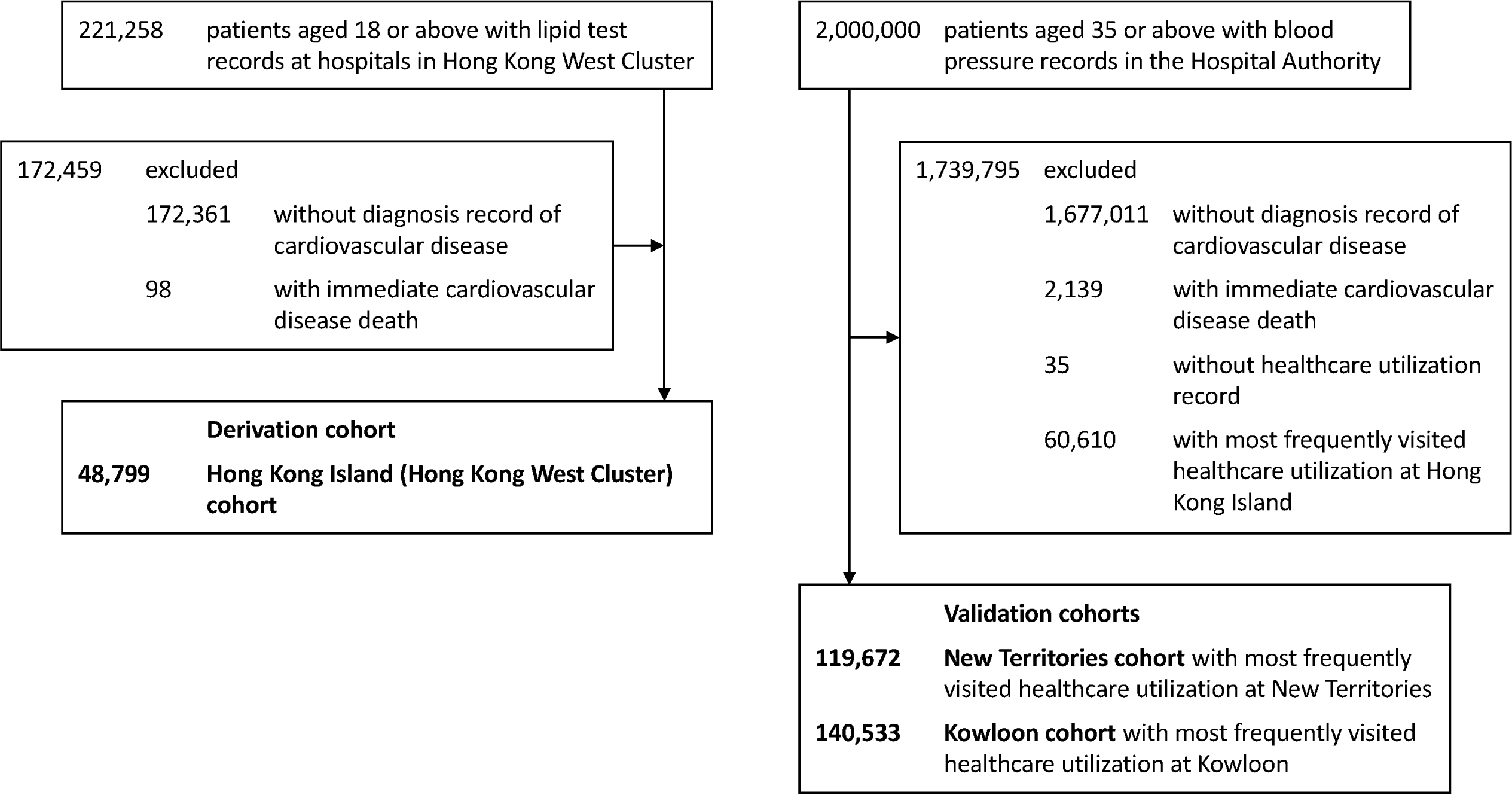
Selection of patients into the study cohorts. N.B. Hong Kong West Cluster is a part of Hong Kong Island.

Each patient was categorised as Hong Kong Island (Hong Kong West Cluster), Kowloon, and New Territories based on the region of their most frequently visited healthcare facility within the study period. Cohort entry date was the date of their first diagnosis of CVD in any inpatient and outpatient setting. Patients were censored at the earliest date of the second record of CVD diagnosis, date of registered death, or study end date (31 December 2019). Patients were excluded from the cohort if they had no diagnosis record of CVD, or died on the same day as the first CVD event.

### Outcomes and risk variables

The outcome was the diagnosis of CVD defined by the International Classification of Diseases, Ninth Revision, Clinical Modification (ICD-9-CM) codes. The outcome was a composite of coronary heart disease, ischaemic or haemorrhagic stroke, peripheral artery disease, and revascularization (Supplementary Table 1). We estimated the incidence of recurrent CVD events for each cohort with reference to the total person-years of each cohort.

The full list of 125 risk variables (Supplementary Table 2) including Commonly known risk factors such as age, sex, lipid profile, blood pressure, haemoglobin A1c, and blood glucose. Of which 15 were mandatory risk variables, were derived based on clinical evidence, statistically strong correlation and data completeness to predict CVD risk. Eight classes of medications including lipid-modifying (fibrates, niacin, cholesterol absorption inhibitors, PCSK9 inhibitors, and statins), antihypertensive, antidiabetic, and antiplatelet drugs were considered interactive drug use options to observe any changes in CVD risk in the model. Diagnoses and procedures were defined by ICD-9-CM codes (Supplementary Table 3), and medication exposure was defined by the British National Formulary (BNF) sections (Supplementary Table 4-5).

### Model derivation

The design of the hybrid statistical-ML model is illustrated in Supplementary Figure 1. Feature selection procedure was first applied to all available risk variables to identify mandatory risk variables for model interpretability. Multivariate imputation with chained equations (MICE) was used to generate one imputed dataset to replace the missing values of clinical laboratory tests.^27^ MICE is a principled method for dealing with missing data and is extremely reliable on high-dimensional datasets with various missing patterns.^28^ For better statistical reliability and clinical utility, risk variables with missing rates below 10% (e.g., clinical laboratory tests) and event rate above 5% (e.g., disease and medication history) were passed for feature selection. We employed a Cox proportional hazards model (CPH) with the least absolute shrinkage and selection operator (LASSO) regularization to shortlist statistically significant (p value<0·05) risk variables.^29,30^ CPH is the most widely used multivariate statistical model for survival analysis.^31,32^ Its regression coefficients can be interpreted as hazard ratios which can be easily understood by clinicians for better decision-making. LASSO is a robust feature selection method. It selects the most representative yet independent set of risk variables, which is reliable when downstream manual prioritisation is required. Mandatory risk variables were also determined based on clinical relevance to ensure the final set of risk variables are comprehensive and relevant to CVD prognosis. Mandatory risk variables were included in the final model as linear covariates.

For better model performance, the measurement and integration of complex effects from all risk variables in the EHR is important for our model. However, real-world EHR data like our cohorts, are highly heterogeneous in form, distribution, and especially completeness. Most ML methods require complete datasets which will cause huge imputation bias in high-dimensional datasets. Compared to other state-of-the-art ML methods e.g., deep learning (neural network), XGBoost is a gradient boosting decision tree method for better dealing with heterogeneous tabular data.^33^ More importantly, it can work with missing values without imputation. Therefore, we used XGBoost in P-CARDIAC to fit a tree-ensembled hazard ratio based on all risk variables (Supplementary Information 2). To cancel out the non-linear distribution bias in the raw output of XGBoost, the raw output hazard ratio was first mapped to discrete percentiles, which was tested to largely benefit model calibration performance. To balance the significance between the XGBoost risk score and other risk variables in the final model, the percentiles are then mapped onto a hinge loss-like function (Supplementary Information 3). The P-CARDIAC full model with all 125 risk variables is a CPH model with ridge regularization regressed on the mandatory risk variables and the XGBoost risk score.^34^ Ridge regularization is widely used as a stabilizer of regression coefficients, which provides reliable estimates of the hazard ratios of the risk variables. For comparison, a CPH model with only the mandatory risk variables was built as a P-CARDIAC basic model.

### Model validation

Internal consistency of model performance was evaluated on the derivation cohort) by 100 repeats of 10-fold cross-validation. Model performance of P-CARDIAC, TRS-2°P and SMART2 was compared using the validation cohorts with 1,000 bootstrap replicates. A high number of repeats were employed to ensure accurate estimation (mean and confidence interval) of model performance statistics.

Calibration performance was assessed graphically by categorising patients into deciles of predicted 10-year CVD risk and plotting mean 10-year predicted risk against observed 10-year risk. The observed 10-year risk was obtained by the Kaplan-Meier method^35^. Means and confidence intervals of Harrell’s C statistic, calibration-in-the-large, and calibration slope were calculated.^36,37^ The calibration slope was the slope of linear regression of the observed risk against the predicted risk of each decile. Recalibration was performed if there was overall overestimation or underestimation observed in the calibration curves.^38^

Decision curve analysis was used to estimate the effect of different treatment options across different threshold risks.^39-41^ This can identify the range of threshold risks where the model has clinical value (with positive net benefit) and the magnitude of the clinical value. The model with higher net benefits across a larger range of threshold risks is the preferred model. We used decision curve analysis to describe and compare the 10-year clinical value of P-CARDIAC, TRS-2°P, and SMART2 on the two validation cohorts. TRS-2°P has proposed the specific 3-year risk regarding different risk scores, and we extrapolated the predicted 3-year risk to 10-year risk by multiplying the ratio of the corresponding Kaplan-Meier estimated risks for each of the two cohorts.

All analysis were conducted using Python (version 3·9·1) with add-on package lifelines.^42^ This study report is in accordance with the TRIPOD statement.^43^ Ethical approval for this study was granted by the Institutional Review Board of the University of Hong Kong/HA Hong Kong West Cluster (UW20-073).

## Results

### Study cohorts

For the derivation cohort, we identified 221,258 patients aged 18 or above with lipid test records between 1 January 2004 and 31 December 2019. We excluded 172,459 patients from the cohort who had no diagnosis record of CVD or died of the first CVD event on the same date. Overall, 48,799 patients were included in the derivation cohort.

For the validation cohorts, we initially identified a cohort of 2 million patients aged 35 or above with blood pressure records in the Hospital Authority between 1 January 2005 and 31 December 2019. We excluded 1,679,150 patients who had no diagnosis record of CVD or died of the first CVD event on the same date. We excluded 60,645 patients without healthcare utilization records or with the most frequently visited healthcare facility at Hong Kong Island. Overall, 119,672 patients were included in the New Territories cohort, and 140,533 patients were included in the Kowloon cohort. A flowchart of patient selection is illustrated in Figure 1.

### Incidence rates of CVD and baseline characteristics

Table 1 shows the event rates of CVD across three cohorts. The event rate per 1000 person-years was 219 to 241, while the median estimated 10-year event rate was 71·7-76·1%, respectively. During a median follow-up of 0·3 to 1·0 year, 55-64% of patients had cardiovascular disease recurrences. Regarding the composition of incident CVD events, coronary heart disease (CHD) was the most common, with composition around 61-65%, of which MI had a ratio of approximately 9-10%. Stroke was the second most common outcome with a ratio of approximately 33-39%. The ratio of peripheral arterial disease (PAD) was around 3-4%.

**Table 1.** Patient characteristics.

|  | Hong Kong Island (Hong Kong West Cluster) | Kowloon | New Territories |
| --- | --- | --- | --- |
| **Participants** | 48,799 | 140,533 | 119,672 |
| **Incident cardiovascular events** | 31,100 (64%) | 80,498 (57%) | 65,687 (55%) |
| Coronary heart disease | 20,167 (65%) | 49,754 (62%) | 39,807 (61%) |
| Myocardial infarction | 3,231 (10%) | 7,341 (9%) | 5,773 (9%) |
| Stroke | 10,394 (33%) | 30,342 (38%) | 25,413 (39%) |
| Peripheral artery disease | 1,102 (4%) | 2,188 (3%) | 1,826 (3%) |
| Revascularization | 4,135 (13%) | 5,396 (7%) | 4,447 (7%) |
| *Fatal events | 964 (3%) | 4,544 (6%) | 3,246 (5%) |
| **Total person-years observed** | 141,829 | 334,053 | 293,269 |
| **Event rate per 1000 person-years** | 219 | 241 | 224 |
| **\*\*Follow-up (years)** | 0.3 (0.0-13.5) | 0.9 (0.0-10.4) | 1.0 (0.0-10.5) |
| **\*\*\*10-year event rate (%)** | 71.7 (71.3-72.2) | 76.1 (75.8-76.5) | 73.3 (72.9-73.7) |

All data in n (%) or median (interquartile range) unless indicated otherwise. All subtypes of incidence events in the Kowloon and New Territories cohorts were significantly different (p value<0.05) compared to the Hong Kong Island (Hong Kong West Cluster) under Chi-square test. Event rate was the incident event divided by total person-years of each cohort.
\*Deaths within 28 days after recurrent cardiovascular event.
\*\*Median (5th/95th percentile).
\*\*\*Mean (95% confidence interval), estimated by Kaplan-Meier method.

All subtypes of incidence events in the derivation cohort had significantly different distribution from the validation cohorts. The proportion of total CVD events was higher. The proportion of CHD, MI, PAD, and revascularization was higher, while the proportion of stroke and fatal events were lower. Table 2 and Supplementary Table 6 showed the baseline characteristics of the risk variables across three cohorts.

**Table 2.** Summary of mandatory risk variables and drug use.

|  | Hong Kong Island (Hong Kong West Cluster) | Kowloon | New Territories |
| --- | --- | --- | --- |
| **General [n (%), or median (interquartile range)]** |  |  |  |
| Age (years) | 69 (59-78) | 73 (63-82) | 71 (61-80) |
| Female | 18,948 (39%) | 61,101 (43%) | 50,187 (42%) |
| Male | 29,851 (61%) | 79,432 (57%) | 69,485 (58%) |
| Accident and emergency visits per year | 0.6 (0.0-0.7) | 0.9 (0.5-1.1) | 0.9 (0.6-1.2) |
| **Clinical laboratory tests [median (interquartile range, proportion of missing data)]** |  |  |  |
| Low-density lipoprotein cholesterol (mmol/L) | 2.5 (1.9-3.1, 0%) | 2.6 (2.0-3.3, 5%) | 2.6 (2.0-3.3, 4%) |
| Neutrophil (10<sup>9</sup>/L) | 4.9 (3.7-6.8, 2%) | 5.3 (3.9-7.8, 3%) | 5.3 (3.9-7.7, 2%) |
| Aspartate transaminase: alanine aminotransferase ratio | 1.1 (0.8-1.6, 1%) | 1.3 (0.9-1.9, 37%) | 1.3 (0.8-2.2, 80%) |
| **Disease and medication history [n (%)]** |  |  |  |
| Statins | 12,801 (26%) | 47,278 (34%) | 42,127 (35%) |
| Hypertension | 30,583 (63%) | 109,374 (78%) | 92,568 (77%) |
| Diabetes | 12,388 (25%) | 43,096 (31%) | 37,217 (31%) |
| Atrial fibrillation | 4,248 (9%) | 13,920 (10%) | 11,251 (9%) |
| Myocardial infarction | 5,361 (11%) | 23,626 (17%) | 18,162 (15%) |
| Angina | 3,548 (7%) | 10,389 (7%) | 7,126 (6%) |
| Revascularization | 6,839 (14%) | 6,199 (4%) | 6,455 (5%) |
| Family history of diabetes | 4,878 (10%) | 17,278 (12%) | 15,613 (13%) |
| **Drug use [n (%)]** |  |  |  |
| Antihypertensive drugs | 38,851 (80%) | 121,287 (86%) | 101,353 (85%) |
| Antidiabetic drugs | 12,995 (27%) | 44,081 (31%) | 37,644 (31%) |
| Antiplatelet drugs | 35,575 (73%) | 116,263 (83%) | 99,051 (83%) |
| Statins | 31,452 (64%) | 90,856 (65%) | 84,260 (70%) |
| Fibrates | 1,201 (2%) | 3,491 (2%) | 2,402 (2%) |
| Niacin | 65 (0%) | 16 (0%) | 20 (0%) |
| PCSK9 inhibitors | 30 (0%) | 22 (0%) | 48 (0%) |
| Cholesterol absorption inhibitors | 666 (1%) | 853 (1%) | 1,102 (1%) |

All risk variables in the Kowloon and New Territories cohorts were significantly different (p value<0.05) compared to the Hong Kong Island (Hong Kong West Cluster) under Chi-square test (categorical risk variables) or in T-test (numerical risk variables). PCSK9 = Proprotein convertase subtilisin/kexin type 9.

### Model derivation

We identified 15 mandatory risk variables and 8 interactive drug use options (Table 3) that were statistically significant and medically coherent for CVD pathogenesis. MICE was conducted once with less than 2% missing rate among the 15 mandatory risk variables. For both the basic and full model, all risk variables were statistically significant (p value<0·05) when compared to those without recurrent CVD. Both models had similar estimates on the linear effects of the risk variables while the basic model’s hazard ratios deviated more than 1 from the full model.

**Table 3.** Adjusted hazard ratios in P-CARDIAC models.

|  | **Basic model**<br>(Mandatory risk variables) |  | **Full model**<br>(Mandatory +<br>Supplementary risk<br>variables) |  |
| --- | --- | --- | --- | --- |
|  | HR (95% CI) | p value | HR (95% CI) | p value |
| **General** |  |  |  |  |
| Age per year | 1.02 (1.01-1.02) | <0.0001 | 1.01 (1.01-1.01) | <0.0001 |
| Female | 0.84 (0.82-0.86) | <0.0001 | 0.86 (0.84-0.88) | <0.0001 |
| Accident and emergency visits per year<br>(prior to incident cardiovascular events) | 1.07 (1.06-1.08) | <0.0001 | 1.06 (1.05-1.07) | <0.0001 |
| **Clinical laboratory tests** |  |  |  |  |
| Low-density lipoprotein cholesterol<br>(mmol/L) | 1.06 (1.05-1.08) | <0.0001 | 1.05 (1.04-1.06) | <0.0001 |
| Neutrophil (10<sup>9</sup>/L) | 1.02 (1.02-1.03) | <0.0001 | 1.02 (1.02-1.02) | <0.0001 |
| Aspartate transaminase: alanine<br>aminotransferase ratio | 1.02 (1.02-1.03) | <0.0001 | 1.02 (1.01-1.02) | <0.0001 |
| **Disease and medication history** |  |  |  |  |
| Statins | 0.84 (0.82-0.87) | <0.0001 | 0.88 (0.85-0.90) | <0.0001 |
| Hypertension | 1.16 (1.13-1.19) | <0.0001 | 1.13 (1.10-1.16) | <0.0001 |
| Diabetes | 1.38 (1.34-1.43) | <0.0001 | 1.30 (1.25-1.35) | <0.0001 |
| Atrial fibrillation | 1.09 (1.05-1.13) | <0.0001 | 1.08 (1.04-1.12) | 0.0001 |
| Myocardial infarction | 2.13 (2.06-2.21) | <0.0001 | 1.71 (1.65-1.78) | <0.0001 |
| Angina | 0.92 (0.88-0.96) | 0.0003 | 0.93 (0.89-0.97) | 0.0022 |
| Revascularization | 0.91 (0.88-0.95) | <0.0001 | 0.93 (0.90-0.96) | <0.0001 |
| Family history of diabetes | 1.37 (1.32-1.43) | <0.0001 | 1.28 (1.23-1.33) | <0.0001 |
| **Drug use** |  |  |  |  |
| Antihypertensive drugs | 0.67 (0.65-0.69) | <0.0001 | 0.77 (0.74-0.79) | <0.0001 |
| Antidiabetic drugs | 0.71 (0.69-0.74) | <0.0001 | 0.77 (0.74-0.80) | <0.0001 |
| Antiplatelet drugs | 0.78 (0.75-0.80) | <0.0001 | 0.85 (0.83-0.87) | <0.0001 |
| Fibrates | 0.78 (0.73-0.84) | <0.0001 | 0.78 (0.73-0.84) | <0.0001 |
| Niacin | 0.53 (0.38-0.75) | 0.0003 | 0.56 (0.40-0.78) | 0.0007 |
| Cholesterol absorption inhibitors | 0.55 (0.49-0.63) | <0.0001 | 0.56 (0.49-0.63) | <0.0001 |
| PCSK9 inhibitors | 0.24 (0.09-0.68) | 0.0066 | 0.25 (0.09-0.69) | 0.0078 |
| Statins | 0.87 (0.85-0.90) | <0.0001 | 0.89 (0.86-0.91) | <0.0001 |
| **XGBoost risk score** |  |  | 1.03 (1.02-1.03) | <0.0001 |

Abbreviations: HR=hazard ratio, CI=confidence interval, PCSK9 = Proprotein convertase subtilisin/kexin type 9.

### Model validation

Validation results on the derivation cohort of P-CARDIAC full model showed satisfying discrimination and calibration performance. The C statistic was 0·69, the calibration slope was 1·00, and the calibration-in-the-large was 0·03. There was slight overestimation across risk deciles. P-CARDIAC basic model showed good discrimination and calibration performance but was inferior to the full model. The C statistic was 0·66, the calibration slope was 0·86, and the calibration-in-the-large was 0·01. There was slight overestimation in high-risk patients and underestimation in low-risk patients. The internal validation results are shown in Figure 2 and Table 4.

**Figure 2.**
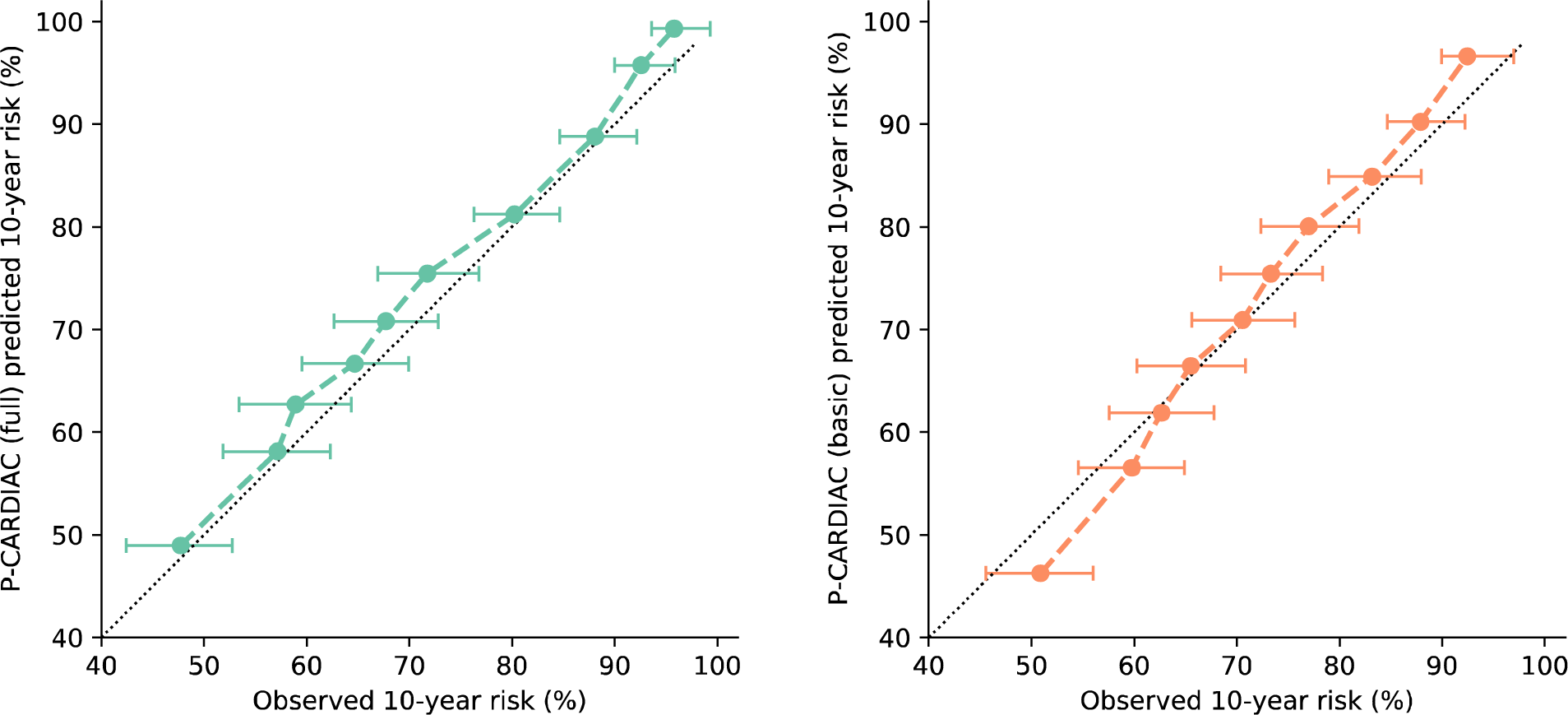
Calibration plots for the P-CARDIAC (full) model in the Hong Kong Island (Hong Kong West Cluster) derivation cohort with 95% Confidence interval. Results were measured from 10-fold cross validation.

**Table 4.** Discrimination and calibration performance of P-CARDIAC on derivation cohort.

|  | Harrell's C statistic | Calibration slope | Calibration-in-the-large |
| --- | --- | --- | --- |
| Basic model | 0.66 (0.66, 0.66) | 0.86 (0.86, 0.86) | 0.01 (0.01, 0.01) |
| Full model | 0.69 (0.69, 0.69) | 1.00 (1.00, 1.00) | 0.03 (0.03, 0.03) |

Harrell's C statistic is a measure of model discrimination with values ranging from 0.5 to 1, i.e., probability of correct ordering for a randomly selected pair of subjects. Calibration slope is a measure of model calibration with target value of 1. Values smaller than 1 indicate overfitting, i.e., too low for low-risk patients and/or too high for high-risk patients. Values greater than 1 indicate underfitting, i.e., too high for low-risk patients and/or too low for high-risk patients. Calibration-in-the-large is a measure of model calibration with target value of 0. Values greater than 0 means the model overestimates risk in general. Values smaller than 0 means the model underestimates risk in general. Results were measured from 100 repeats of 10-fold cross validation.

Internal validation of the P-CARDIAC full model across validation cohorts showed good discrimination and calibration performance. The C statistic for the Kowloon and New Territories cohorts were 0·62 and 0·64, the calibration slope was 0·75 and 0·93, and the calibration-in-the-large was 0·04 and 0·01, respectively. There was overestimation for high-risk patients (predicted 10-year risk greater than 80%) for the Kowloon cohort. There was overestimation on all patients for the New Territories cohort which was largely mitigated by recalibration (Supplementary Figure 2). The P-CARDIAC basic model showed good discrimination and calibration performance but was inferior to the full model. The C statistic for Kowloon and New Territories cohorts were 0·60 and 0·62, the calibration slope was 0·66 and 0·75, and the calibration-in-the-large was 0·01 and 0·03, respectively. There was overestimation in high-risk patients and underestimation in low-risk patients for both cohorts which could not be mitigated by recalibration. Validation of both TRS-2°P and SMART2 risk scores underperformed regarding discrimination and risk stratification performance. The C statistic was lower than 0·55 for both validation cohorts. The validation results are summarised in Figure 3 and Tables 5-7.

**Figure 3.**
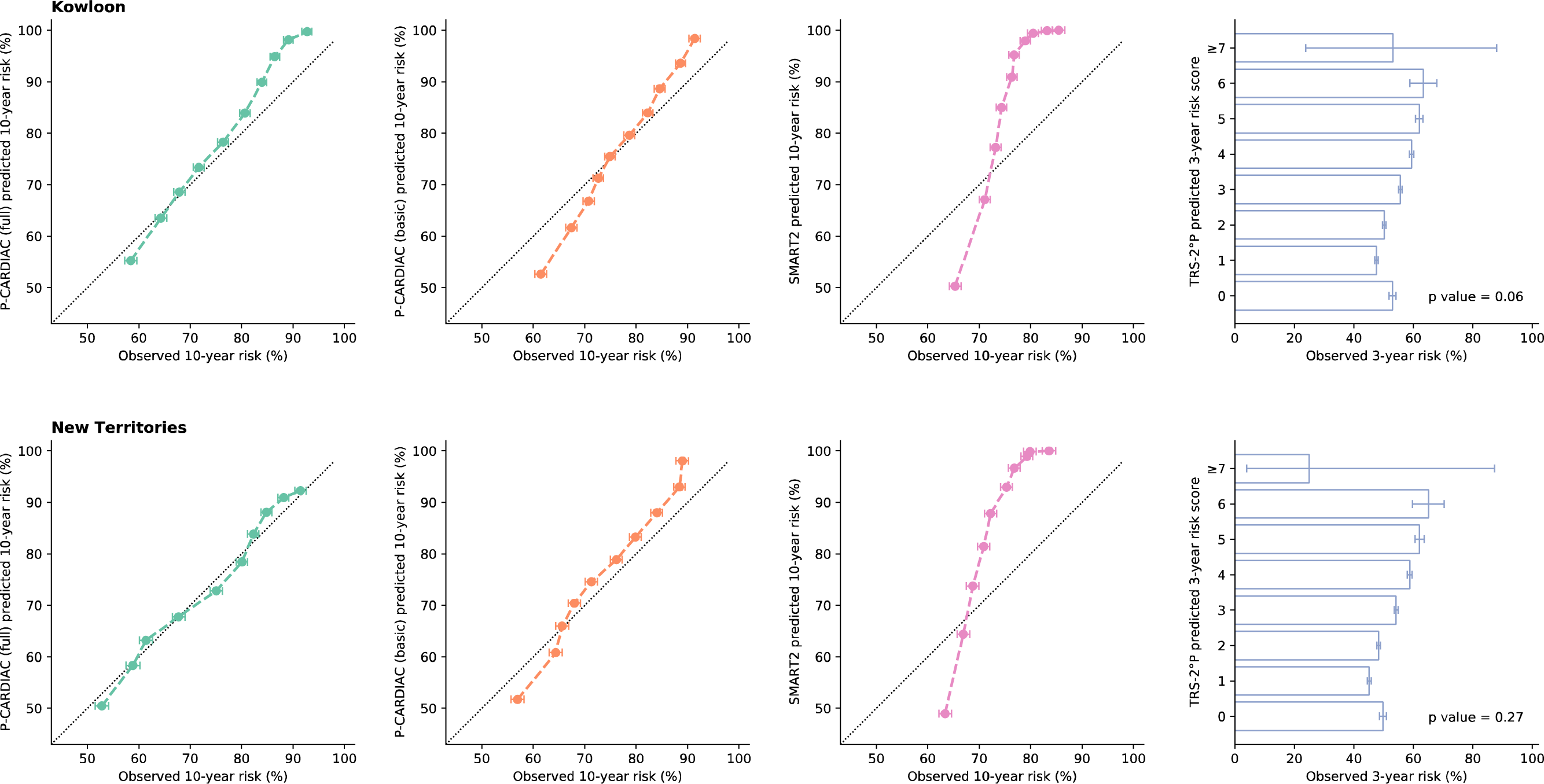
Calibration plots on validation cohorts. Error bar was 95% confidence interval. P values on the right two figures were Mann-Kendall test for significance of monotonic trend. P value larger than 0.05 indicated no significant sign of increasing or decreasing trend in observed risk when predicted risk score increases. Result of P-CARDIAC (full) validated on New Territories cohort (lower left) was after recalibration.

**Table 5.** Mean (95% CI) of Harrell’s C statistic on validation cohorts.

|  | P-CARDIAC (full) | P-CARDIAC (basic) | SMART2 | TRS-2°P |
| --- | --- | --- | --- | --- |
| Kowloon | 0.62 (0.62, 0.62) | 0.60 (0.60, 0.60) | 0.55 (0.55, 0.55) | 0.53 (0.53, 0.53) |
| New Territories | 0.64 (0.64, 0.64) | 0.62 (0.62, 0.62) | 0.55 (0.55, 0.55) | 0.54 (0.54, 0.54) |

A measure of model discrimination with values ranging from 0.5 to 1, i.e., probability of correct ordering for a randomly selected pair of subjects. CI=confidence interval. Values were measured from 1000 bootstrap replicates.

**Table 6.** Mean (95% CI) of calibration slope on validation cohorts.

|  | P-CARDIAC (full) | P-CARDIAC (basic) | SMART2 |
| --- | --- | --- | --- |
| Kowloon | 0.75 (0.74, 0.75) | 0.66 (0.66, 0.66) | 0.38 (0.38, 0.38) |
| New Territories* | 0.93 (0.93, 0.93) | 0.75 (0.75, 0.75) | 0.39 (0.39, 0.39) |

A measure of model calibration with target value of 1. Values smaller than 1 indicate overfitting, i.e., too low for low-risk patients and/or too high for high-risk patients. Values greater than 1 indicate underfitting, i.e., too high for low-risk patients and/or too low for high-risk patients. CI=confidence interval. Values were measured from 1000 bootstrap replicates. \*After recalibration.

**Table 7.** Mean (95% CI) of calibration-in-the-large on validation cohorts.

|  | P-CARDIAC (full) | P-CARDIAC (basic) | SMART2 |
| --- | --- | --- | --- |
| Kowloon | 0.04 (0.04, 0.04) | 0.01 (0.01, 0.01) | 0.10 (0.10, 0.10) |
| New Territories* | 0.01 (0.01, 0.01) | 0.03 (0.03, 0.03) | 0.11 (0.11, 0.11) |

A measure of model calibration with target value of 0. Values greater than 0 means the model overestimates risk in general. Values smaller than 0 means the model underestimates risk in general. CI=confidence interval. Values were measured from 1000 bootstrap replicates. \*After recalibration.

In summary, P-CARDIAC showed great performance on the three derivation and validation cohorts. The full model had better performance than the basic model as it accurately accounted for the nonlinear effects and the effects from supplementary risk variables. On the other hand, TRS-2°P and SMART2 underperformed when adapted to the two cohorts for Chinese populations.

### Clinical utility

Decision curve analysis of the two validation cohorts was similar (Figure 4). P-CARDIAC full model performed better than the P-CARDIAC basic model. Both P-CARDIAC models had similar and greater net benefits across a larger range of threshold risks compared with the treat all strategy, TRS-2°P, and SMART2. P-CARDIAC had clinical values for decision-making when the threshold risk was under 90%.

**Figure 4.**
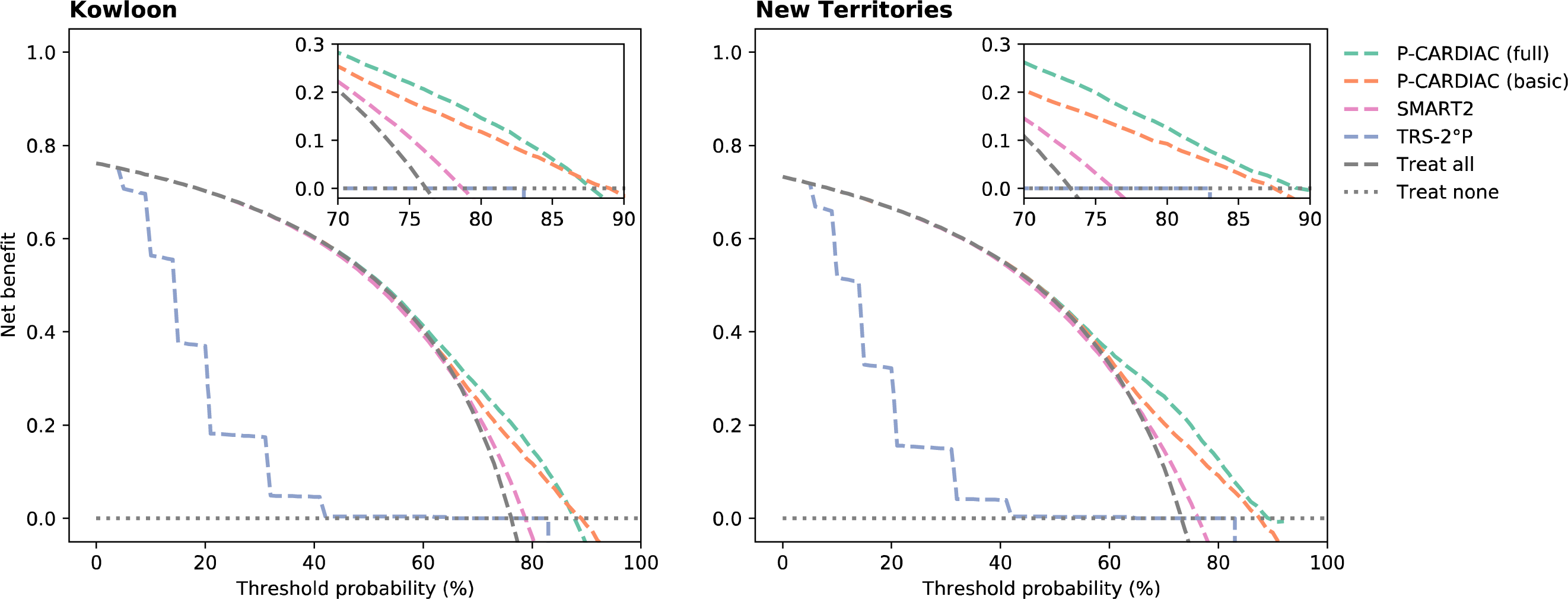
Decision curves on validation cohorts. The threshold probability was the predicted 10-year cardiovascular disease recurrence risk. Result of P-CARDIAC (full) validated on New Territories cohort (right) was after recalibration.

### Website design

The website interface at p-cardiac.com was designed to be flexible and interactive (see Supplementary Information 4 for example screenshots). Users can input up to 15 risk variables in the mandatory field for a quick evaluation of CVD risk. More than 100 risk variables can be further inputted in the supplementary field for a more comprehensive evaluation. The more risk variables submitted in the supplementary field, the more accurate the prediction. Furthermore, the drug use risk variables were designed as interactive selection options, where up to 8 types of drug classes could be selected for evaluation of potential synergetic treatment effects to guide possible treatment plans.

## Discussion

To the best of our knowledge, this is the first model to predict recurrent CVD events in a Chinese population from a large contemporary Chinese cohort using ML technique. P-CARDIAC demonstrated reliable performance of recurrent CVD risk prediction in 10 years on three derivation and validation cohorts. We demonstrated that P-CARDIAC models have better performance in risk prediction than existing CVD risk scores such as TRS-2°P and SMART2 that were developed on western populations. Our results also demonstrate that the P-CARDIAC full model has superior performance to the basic model.

In addition, the effects of concurrent drug use are often neglected in existing CVD risk scores. In this study, we included exposures of various drug classes as interactive covariates in the model to evaluate their bias-mitigated, risk stratified, and Chinese-specific treatment effects. Among the 8 drug classes included in the interactive covariates, all classes had hazard ratios lower than 1 whilst PSCK9 inhibitors had the lowest. This observation indicates that drug treatment with indications for risk variable CVD such as lipid-modifying drugs, antihypertensive, and antidiabetic drugs all have a beneficial effect on reducing CVD risk. In addition, our model also considers prior statin use for primary prevention prior to the first CVD event. We found that patients who received statins as primary prevention prior to the first CVD event had a lower risk of recurrent CVD events, independent of whether they continued statin therapy. We believe P-CARDIAC is the first risk prediction model to include these risk variables in CVD risk prediction, highlighting the novelty of our approach.

P-CARDIAC was developed using hybrid statistical-machine learning algorithms, which is novel in the field of CVD risk prediction. To facilitate efficient clinical management, comprehensive electronic systems were developed, thus providing sizable clinical data for better development of computational models. However, as the pool of covariates becomes increasingly larger, there is a dilemma in the development of medical prediction models, where it may be challenging to balance interpretability and performance. Traditional prediction tools rely on the linear combinations of a selected pool of small number of covariates, which are easily interpreted, but do not consider the massive nonlinear effects and often lack accuracy. On the other hand, in recent years many Machine-Learning (ML) and deep learning methods have emerged that takes into consideration the complex relationships of all massive covariates to yield high accuracy. However, since these models lack linear representations of the covariates, the effects of the risk variables are uncertain and unclear^44^. Therefore, the ML approach is described as the “black box approach”. Our proposed methodology adopts the traditional approach by selecting a pool of clinically relevant covariates using statistical methods, then considers the large number of covariates and their complex effects using the ML method as another non-interfering component for better model fit. We used XGBoost as the ML method. XGBoost is a tree-based ensemble method that does not require complete values in the large pool of covariates which circumvents the potential imputation bias. This novel hybrid method showed significantly better performance than the traditional statistical method by comprehensively considering a large pool of covariates, including commonly known risk factors, such as blood pressure, haemoglobin A1c, blood glucose, and lipid profilewhere its interpretability is still evident. The novel hybrid method is customisable and can be used for other studies.

This study has limitations. First, P-CARDIAC was developed using real-world data and any change in clinical practice in future may result in changes to the predicted recurrent CVD risk among patients. The advantage of the ML approach is that recalibration and fine-tuning the model can be done as more data is accrued. Therefore, the model can be calibrated periodically to account for any changes in clinical practice. Second, P-CARDIAC was developed based on a population of predominantly Chinese, hence recalibration is needed for use in populations of other ethnicities. Third, manual input of more than 100 risk variables is time consuming and not practical in fast-paced clinical settings. Therefore, we aim to automate the process of data entry by leveraging the readily available EHR for clinical management to provide timely risk estimation. Last, P-CARDIAC serves as a risk stratification tool to better utilise healthcare resources rather than a diagnostic tool, thus, a composite risk score was given for a spectrum of CVD diseases rather than a score for each specific disease. The advanced technologies currently available enables the harnessing of the power of Big Data. However, we believe that the empathy of healthcare providers and their connection with patients which influences the best decision on care will not be replaced by AI in the near future.

## Conclusions

We developed and validated P-CARDIAC, a new CVD risk prediction model for recurrent CVD events among Chinese adults with established CVD. Compared to TRS-2°P and SMART2, P-CARDIAC was able to identify unique patterns of Chinese patients with established CVD with good performance. The consideration of treatment effects of various drug use could also guide improved and individualised secondary prevention. We anticipate that P-CARDIAC will have clinical application in a variety of settings, including primary care where real-world data will provide guidance for early intervention of lifestyle changes and potentially promote medication adherence to prevent recurrent CVD events, thus reducing the related healthcare burden.

## Clinical Perspectives

We developed a first CVD risk prediction model for secondary prevention, the Personalized CARdiovascular DIsease risk Assessment for Chinese (P-CARDIAC), using the ML technique with more than 120 risk factors among Chinese population in Hong Kong. The application of ML facilitates a better model performance with large datasets enables cost-effective decision-making through a time variation effect. P-CARDIAC was validated by its derived cohort and other two independent cohorts in Hong Kong. The validation results showed satisfying discrimination and calibration with over 0·6 C-statistic. Our model also showed better performance than other popular recurrent risk models, such as TRS-2°P and SMART2, among Chinese population with established CVD events. The synergetic effect of the treatment with time-variation has always been neglected in risk score calculation.

Particularly, P-CARDIAC included the treatment exposures of various drug classes as interactive covariates in the model to re-evaluate the recurrent risk. Our study indicated that common CVD medication, such as lipid-modifying drugs, antihypertensive, and antidiabetic drugs had a beneficial effect on reducing risk. We particularly considered prior statins as primary prevention for the first CVD episode which could lower the recurrent risk. P-CARDIAC is an innovative model with the application of hybrid statistical-machine learning algorithms to propose and facilitate further prediction model development with more complex relationships involving massive risk factors and populations. We believe P-CARDIAC has the potential to provide guidance in the early intervention treatment of high-risk recurrent CVD patients, such as lifestyle changes and medication compliance, in the future.

## Data Availability

All data produced in the present study are available upon reasonable request to the authors

## Abbreviations list

CVD: Cardiovascular Disease
P-CARDIAC: Personalized CARdiovascular DIsease risk Assessment for Chinese
TRS-2°P: Thrombolysis in Myocardial Infarction (TIMI) Risk Score for Secondary Prevention
SMART2: Secondary Manifestations of ARTerial disease
ML: Machine-Learning
EHR: Electronic Health Records
HA: Hospital Authority
ICD-9-CM: Ninth Revision, Clinical Modification
BNF: British National Formulary
MICE: Multivariate imputation with chained equations
CPH: Cox proportional hazards model
LASSO: Least Absolute Shrinkage and Selection Operator
CHD: Coronary Heart Disease
PAD: Peripheral Arterial Disease
MI: Myocardial Infarction

## Acknowledgement

We thank Ms. Lisa Lam for proof editing.

## Data sharing statement

Data will not be available for others as the data custodians have not given permission.

