## Supplementary materials for "Development and validation of risk prediction model for recurrent cardiovascular events among Chinese: P-CARDIAC model"

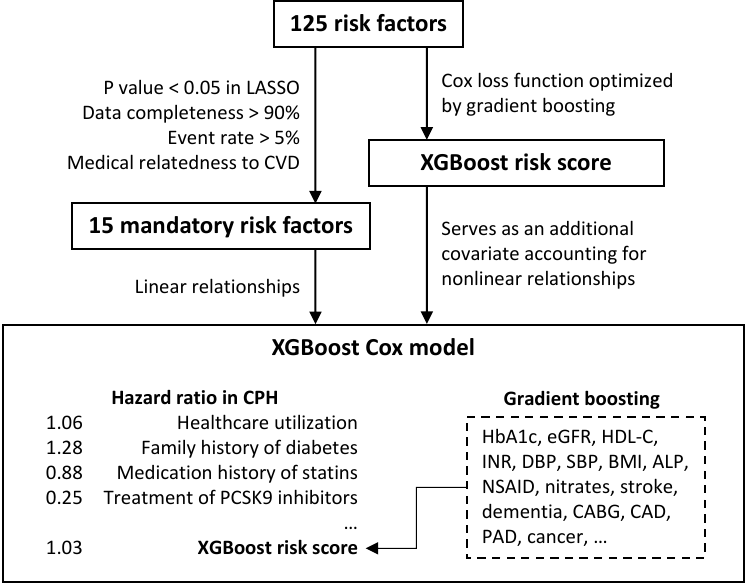

**Supplementary Figure 1. Algorithm design of XGBoost Cox model used in P-CARDIAC full model.** PCSK9 = Proprotein convertase subtilisin/kexin type 9.

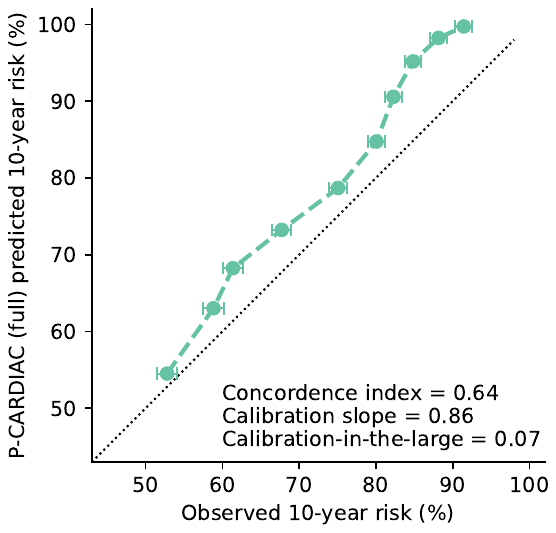

**Supplementary Figure 2.** Validation results of P-CARDIAC (full) on New Territories before recalibration.

**Supplementary Table 1. Definition of cardiovascular disease**

| **Diagnosis** | **ICD-9** |
| --- | --- |
| Peripheral artery disease | 440, 443.9 |
| Coronary heart disease | 410-414, 429.2, V45.81 |
| Myocardial infarction | 410 |
| Stroke | 430, 431, 432, 433.01, 433.11, 433.21, 433.31, 433.81, 433.91, 434, 435, 436, 437.0, 437.1 |
| **Procedure** |  |
| Revascularization | 36.01-36.20 |

**Supplementary Table 2. Summary of included variables**

| Categories (number of covariates) | Risk variables |
| --- | --- |
| **Demographic factors (2)** | age*, sex* |
| **Family history of disease (2)** | diabetes*, cardiovascular disease |
| **Healthcare utilization (3)** | accident and emergency visits per year*, inpatient visits per year, outpatient visits per year |
| **Clinical laboratory tests (39)** | aspartate transaminase*, alanine aminotransferase*, low-density lipoprotein cholesterol*, neutrophil*, hemoglobin A1c, creatine kinase (total), prothrombin time, potassium (serum), estimated glomerular filtration rate, triglycerides, basophil, arterial partial pressure of oxygen, albumin, international normalized ratio, diastolic blood pressure, bicarbonate (serum), glucose (fasting), erythrocyte sedimentation rate, free thyroxine, troponin I, bilirubin (total), C-reactive protein, total cholesterol, blood pH, systolic blood pressure, thyroid stimulating hormone, lymphocyte, creatinine (serum), platelet, red blood cell, high-density lipoprotein cholesterol, body mass index, calcium (serum), white blood cell, alkaline phosphatase, sodium (serum), eosinophil, hemoglobin, monocyte |
| **Medication history (prior to incident CVD event) (27)** | statins*, antihypertensive drugs, antidiabetic drugs, antiplatelet drugs, non-steroidal anti-inflammatory drugs, corticosteroids, proton-pump inhibitors, H2-receptor antagonists, anticoagulants, nicotine replacement therapy, antiarrhythmic drugs, antithyroid drugs, oestrogen, psychotropic drugs, cardiac glycosides, nitrates, thyroid hormones, testosterone, fibrates, niacin, PCSK9 inhibitors, cholesterol absorption inhibitors, Vytorin, bile acid sequestrants, omega-3 fatty acids, other non-statin lipid-modifying drugs, count of medication |
| **Disease history (44)** | myocardial infarction*, angina*, revascularization*, atrial fibrillation*, hypertension*, diabetes*, congestive heart failure, stroke, thyroid disease, arrhythmia and conduction disorders, obesity, coronary heart disease, hypothyroidism, cardiac wall/valve/shunt replacement/repairment, oxygen therapy/ventilator/intubation, asthma, injury and poisoning, alcohol user, dyslipidemia, cardiomyopathy, Parkinson’s disease, defibrillator insertion, major organ bleeding, severe mental illness, dementia, pacemaker implantation, liver disease, chronic obstructive pulmonary disease, cancer, rheumatoid arthritis, renal disease, smoker, chronic kidney disease, muscle pain or myopathy or rhabdomyolysis, dialysis, Creutzfeldt-Jakob disease, cardioversion, nephrotic syndrome, coronary artery bypass graft, systemic lupus erythematosus, heart transplantation, peripheral artery disease, migraine, Down’s syndrome |
| **Drug use (after incident CVD event) (8)** | antihypertensive drugs, antidiabetic drugs, antiplatelet drugs, statins, fibrates, niacin, PCSK9 inhibitors, cholesterol absorption inhibitors |

*Mandatory risk variables. PCSK9 = Proprotein convertase subtilisin/kexin type 9. H2 = histamine type 2.

**Supplementary Table 3. Disease list.**

| **Disease/Symptoms** | **ICD-9-CM code** |
| --- | --- |
| Atrial fibrillation | 427.3 |
| Renal disease | 403.01, 403.11, 403.91, 404.02, 404.03, 404.12, 404.13, 404.92, 404.93, 580, 582, 583.0-583.7, 585-587, 588.0, 589, 590, 593.0-593.2, 593.6, 593.8, 593.9, 599.7, 753.0-753.4, 966.1, V42.0, V45.1, V56 |
| Chronic kidney disease | 585 |
| Dialysis | 585.9, V56.0, V56.8, 39.95 |
| Congestive heart failure | 428 |
| Diabetes | 250 |
| Down’s syndrome | 758.0 |
| Hypertension | 401-405 |
| Arrhythmia and conduction disorders | 426, 427 |
| Cardiomyopathy | 425 |
| Angina | 413 |
| Coronary artery bypass graft | 414.04, V45.81 |
| Myocardial infarction | 410 |
| Dyslipidaemia | 272 |
| Thyroid disease | 240-244 |
| Liver disease | 570-573 |
| Migraine | 346 |
| Nephrotic syndrome | 581 |
| Rheumatoid arthritis | 446.5, 710.0-710.4, 714.0-714.3, 725 |
| Several mental illnesses | 290-319 |
| Systemic lupus erythematosus | 710.0 |
| Obesity | 278 |
| Dementia | 290, 291, 292.82, 294, 331 |
| Chronic obstructive pulmonary disease | 490-492, 494, 496 |
| Asthma | 493 |
| Alcohol use | 265.2, 291, 303, 305.0, 357.5, 425.5, 535.3, 571.0- 571.3, 980, V11.3 |
| Smoker | 305.1, V15.82, V15.83, 649.0 |
| Cancer | 140-209, 230-239 |
| Pacemaker implantation | 37.7, 37.8 |
| Defibrillator insertion | 37.94-37.98 |
| Cardioversion | 99.61 |
| Cardiac wall/valve/shunt replacement/repairment | 39.0-39.2 |
| Echocardiography | 37.28 |
| Heart transplantation | 37.51 |
| Oxygen therapy/ventilator/intubation | 00.49, 93.90, 96.01-96.05, 96.7 |
| Erectile dysfunction | 607.84 |
| Major organ bleeding | 578.0, 578.1 |
| Muscle pain, myopathy, or rhabdomyolysis | 728.8, 729.9, 791.3, 781.99 |
| Injury and poisoning | 800-989 |
| Parkinson’s disease | 332 |
| Huntington’s disease | 333.4 |
| Mild cognitive impairment | 331.83 |
| Memory loss | 780.93 |
| Creutzfeldt-Jakob disease | 046.1 |
| Hypothyroidism | 243-244 |

**Supplementary Table 4. Drug list**

| **Drug class** | **BNF chapter** |
| --- | --- |
| Corticosteroids | 1.5.2, 1.7.2, 3.2, 6.3, 8.2.2,10.1.2, 11.4.1, 13.4 |
| H2-receptor antagonists | 1.3.1 |
| Proton-pump inhibitors | 1.3.5 |
| Cardiac glycosides | 2.1.1 |
| Anti-arrhythmic drugs | 2.3.2 |
| Psychotropic drugs | 4.1, 4.2, 4.3, 4.4 |
| Antihypertensive drugs | 2.2, 2.4, 2.5.1, 2.5.2, 2.5.4, 2.5.5, 2.6.2 |
| Nitrates | 2.6.1 |
| Anticoagulants | 2.8.1, 2.8.2 |
| Antiplatelet drugs | 2.9 |
| Antidiabetic drugs | 6.1.1.1, 6.1.1.2, 6.1.2.1, 6.1.2.2, 6.1.2.3 |
| Lipid-modifying drugs* | 2.12 |
| Nicotine replacement therapy | 4.10.2 |
| Oestrogen | 6.4.1 |
| Testosterone | 6.4.2 |
| Non-steroidal anti-inflammatory drugs | 10.1.1 |
| Thyroid hormones | 6.2.1 |
| Antithyroid drugs | 6.2.2 |

*Further distinguish the subclasses based on drug names. H2 = histamine type 2.

**Supplementary Table 5. Lipid-modifying drugs subclasses**

| **Subclass** | **Drug name** |
| --- | --- |
| Statins | Atorvastatin, Fluvastatin, Lovastatin, Pravastatin, Rosuvastatin, Simvastatin |
| Fibrates | Bezafibrate, Clofibrate, Fenofibrate, Gemfibrozil |
| Niacin | Nicotinic acid, Nicotinate, Tredaptive, Acipimox |
| PCSK9 inhibitors | Alirocumab, Evolocumab |
| Cholesterol absorption inhibitors | Ezetimibe |
| Bile acid sequestrants | Cholestyramine |
| Omega-3 fatty acids | Maxepa |
| Vytorin | Vytorin |
| Others | Benfluorex, Probucol |

PCSK9 = Proprotein convertase subtilisin/kexin type 9.

| **Supplementary Table 6. Summary of supplementary variables** | | | | | | |
| --- | --- | --- | --- | --- | --- | --- |
|  | **Hong Kong Island (Hong Kong West Cluster)** | | **Kowloon** | | **New Territories** | |
| **Clinical laboratory tests [median (interquartile range, proportion of missing data)]** | | | | | | |
| Aspartate transaminase (IU/L) | 25.0 | (20.0-33.0, 1%) | 24.0 | (18.0-35.0, 37%) | 27.0 | (20.0-45.0, 80%) |
| Alanine aminotransferase (IU/L) | 23.0 | (16.0-34.0, 1%) | 19.0 | (14.0-28.9, 0%) | 20.0 | (14.0-30.0, 0%) |
| Haemoglobin A1c (%) | 6.1 | (5.7-6.9, 24%) | 6.1 | (5.7-6.8, 16%) | 6.1 | (5.7-6.8, 14%) |
| Creatine kinase (IU/L) | 109.0 | (70.0-196.0, 14%) | 115.0 | (71.0-212.0, 11%) | 113.0 | (72.0-201.1, 10%) |
| Prothrombin time (second) | 11.7 | (11.0-12.5, 7%) | 11.6 | (10.8-12.5, 7%) | 11.4 | (10.7-12.2, 8%) |
| Potassium (mmol/L) | 4.0 | (3.7-4.3, 0%) | 4.0 | (3.7-4.4, 0%) | 4.0 | (3.6-4.3, 0%) |
| Estimated glomerular filtration rate (mL/min/1.73 m^2) | 69.6 | (52.4-84.0, 33%) | 70.0 | (53.5-85.0, 24%) | 73.0 | (57.0-87.0, 29%) |
| Triglycerides (mmol/L) | 1.2 | (0.9-1.6, 0%) | 1.2 | (0.9-1.7, 4%) | 1.2 | (0.9-1.7, 4%) |
| Basophil (10^9/L) | 0.0 | (0.0-0.0, 2%) | 0.0 | (0.0-0.0, 3%) | 0.0 | (0.0-0.1, 2%) |
| Arterial partial pressure of oxygen (kPa) | 11.5 | (6.8-16.1, 51%) | 8.8 | (4.6-14.3, 37%) | 9.0 | (4.7-14.0, 43%) |
| Albumin (g/L) | 41.0 | (37.0-44.0, 1%) | 39.0 | (35.0-42.0, 0%) | 39.4 | (36.0-42.3, 0%) |
| International normalized ratio | 1.0 | (1.0-1.1, 7%) | 1.0 | (1.0-1.1, 7%) | 1.0 | (1.0-1.1, 8%)* |
| Diastolic blood pressure (mmHg) | 73.0 | (65.0-82.0, 46%) | 74.0 | (66.0-84.0, 0%) | 75.0 | (67.0-85.0, 0%) |
| Bicarbonate (mmol/L) | 23.9 | (21.2-26.4, 5%) | 24.0 | (21.0-26.6, 31%) | 23.9 | (21.0-26.5, 39%) |
| Glucose (mmol/L) | 5.7 | (5.1-6.8, 4%) | 5.7 | (5.1-6.9, 4%) | 5.7 | (5.1-6.9, 3%) |
| Erythrocyte sedimentation rate (mm/hr) | 45.0 | (20.0-85.0, 54%) | 37.0 | (19.0-69.0, 50%) | 34.0 | (16.0-65.0, 48%) |
| Free thyroxine (pmol/L) | 16.0 | (13.9-18.1, 51%) | 14.3 | (12.5-16.5, 56%) | 14.8 | (12.8-17.2, 56%) |
| Troponin I (ng/mL) | 0.0 | (0.0-0.1, 53%) | 0.0 | (0.0-0.1, 41%) | 0.0 | (0.0-0.1, 54%) |
| Bilirubin (umol/L) | 9.2 | (7.0-13.0, 1%) | 10.0 | (7.0-14.2, 0%) | 10.0 | (7.0-14.0, 0%) |
| C-reactive protein (mg/dL) | 1.3 | (0.3-5.8, 53%) | 2.0 | (0.4-7.5, 38%) | 1.2 | (0.3-5.7, 36%) |
| Total cholesterol (mmol/L) | 4.3 | (3.6-5.1, 0%) | 4.5 | (3.8-5.3, 4%) | 4.5 | (3.8-5.3, 4%) |
| Blood pH | 7.4 | (7.4-7.5, 47%) | 7.4 | (7.4-7.4, 35%) | 7.4 | (7.4-7.4, 43%) |
| Systolic blood pressure (mmHg) | 135.0 | (122.0-149.0, 46%) | 139.0 | (125.0-155.0, 0%) | 138.0 | (124.0-154.0, 0%) |
| Thyroid stimulating hormone (mIU/L) | 1.3 | (0.9-2.1, 30%) | 1.3 | (0.8-2.1, 15%) | 1.4 | (0.9-2.1, 15%) |
| Lymphocyte (10^9/L) | 1.6 | (1.2-2.1, 2%) | 1.5 | (1.1-2.1, 3%) | 1.6 | (1.1-2.1, 2%)* |
| Creatinine (umol/L) | 88.0 | (73.0-109.0, 0%) | 86.0 | (70.0-109.0, 0%)* | 84.0 | (69.0-104.0, 0%) |
| Platelet (10^9/L) | 223.0 | (184.0-268.0, 2%) | 222.0 | (181.0-269.0, 1%)* | 222.0 | (182.0-268.0, 1%)* |
| Red blood cell (10^12/L) | 4.4 | (4.0-4.8, 2%) | 4.4 | (3.9-4.8, 1%) | 4.4 | (4.0-4.8, 1%) |
| High-density lipoprotein cholesterol (mmol/L) | 1.2 | (0.9-1.4, 0%) | 1.2 | (1.0-1.5, 5%) | 1.2 | (1.0-1.5, 4%) |
| Body mass index (kg/m^2) | 24.7 | (22.2-27.3, 62%) | NA | (NA, 100%) | NA | (NA, 100%) |
| Calcium (mmol/L) | 2.3 | (2.2-2.4, 13%) | 2.3 | (2.2-2.4, 5%) | 2.3 | (2.2-2.4, 4%) |
| White blood cell (10^9/L) | 7.4 | (6.0-9.4, 2%) | 8.0 | (6.4-10.4, 1%) | 7.9 | (6.3-10.2, 1%) |
| Alkaline phosphatase (IU/L) | 73.6 | (61.0-90.0, 1%) | 75.0 | (62.0-92.0, 0%) | 74.0 | (61.0-91.0, 0%) |
| Sodium (mmol/L) | 141.0 | (138.0-143.0, 0%) | 139.8 | (137.0-141.9, 0%) | 139.9 | (137.3-141.6, 0%) |
| Eosinophil (10^9/L) | 0.1 | (0.1-0.2, 2%) | 0.1 | (0.0-0.2, 3%) | 0.1 | (0.0-0.2, 2%) |
| Haemoglobin (g/dL) | 13.4 | (12.1-14.5, 2%) | 13.1 | (11.7-14.3, 1%) | 13.3 | (11.9-14.4, 1%) |
| Monocyte (10^9/L) | 0.4 | (0.3-0.6, 2%) | 0.5 | (0.4-0.7, 3%) | 0.5 | (0.4-0.7, 2%) |
| **Disease history [n (%)]** |  |  |  |  |  |  |
| Congestive heart failure | 3,726 | (8%) | 13,824 | (10%) | 10,715 | (9%) |
| Stroke | 16,985 | (35%) | 62,743 | (45%) | 54,163 | (45%) |
| Thyroid disease | 1,019 | (2%) | 3,455 | (2%) | 2,720 | (2%) |
| Arrhythmia and conduction disorders | 5,956 | (12%) | 21,115 | (15%) | 16,378 | (14%) |
| Obesity | 633 | (1%) | 3,460 | (2%) | 3,202 | (3%) |
| Coronary heart disease | 30,662 | (63%) | 76,562 | (54%) | 64,128 | (54%) |
| Hypothyroidism | 433 | (1%) | 1,695 | (1%) | 1,421 | (1%) |
| Cardiac wall/valve/shunt replacement/repairment | 205 | (0%) | 322 | (0%) | 224 | (0%) |
| Oxygen therapy/ventilator/intubation | 1,359 | (3%) | 8,518 | (6%) | 5,589 | (5%) |
| Asthma | 740 | (2%) | 2,413 | (2%) | 1,765 | (1%) |
| Injury and poisoning | 5,164 | (11%) | 20,980 | (15%) | 17,579 | (15%) |
| Alcohol user | 313 | (1%) | 1,008 | (1%) | 914 | (1%) |
| Dyslipidaemia | 7,047 | (14%) | 34,258 | (24%) | 28,764 | (24%) |
| Cardiomyopathy | 407 | (1%) | 661 | (0%) | 653 | (1%) |
| Parkinson’s disease | 303 | (1%) | 1,134 | (1%) | 880 | (1%) |
| Defibrillator insertion | 154 | (0%) | 146 | (0%) | 68 | (0%) |
| Major organ bleeding | 187 | (0%) | 727 | (1%) | 604 | (1%) |
| Severe mental illness | 3,929 | (8%) | 16,181 | (12%) | 13,874 | (12%) |
| Dementia | 1,810 | (4%) | 9,041 | (6%) | 7,083 | (6%) |
| Pacemaker implantation | 635 | (1%) | 1,296 | (1%) | 1,241 | (1%) |
| Liver disease | 1,187 | (2%) | 5,385 | (4%) | 3,874 | (3%) |
| Chronic obstructive pulmonary disease | 1,527 | (3%) | 7,240 | (5%) | 5,793 | (5%) |
| Cancer | 3,328 | (7%) | 9,991 | (7%) | 7,517 | (6%) |
| Rheumatoid arthritis | 334 | (1%) | 867 | (1%) | 659 | (1%) |
| Renal disease | 3,268 | (7%) | 12,455 | (9%) | 9,425 | (8%) |
| Smoker | 274 | (1%) | 2,776 | (2%) | 895 | (1%) |
| Chronic kidney disease | 1,798 | (4%) | 6,259 | (4%) | 4,450 | (4%) |
| Muscle pain or myopathy or rhabdomyolysis | 137 | (0%) | 673 | (0%) | 449 | (0%) |
| Dialysis | 1,357 | (3%) | 5,215 | (4%) | 3,479 | (3%) |
| Creutzfeldt-Jakob disease | 3 | (0%) | 2 | (0%) | 1 | (0%) |
| Cardioversion | 31 | (0%) | 3 | (0%) | 4 | (0%) |
| Nephrotic syndrome | 189 | (0%) | 767 | (1%) | 562 | (0%) |
| Coronary artery bypass graft | 7 | (0%) | 17 | (0%) | 2 | (0%) |
| Systemic lupus erythematosus | 117 | (0%) | 169 | (0%) | 133 | (0%) |
| Heart transplantation | 5 | (0%) | 10 | (0%) | 5 | (0%) |
| Peripheral artery disease | 1,475 | (3%) | 3,244 | (2%) | 2,770 | (2%) |
| Migraine | 51 | (0%) | 143 | (0%) | 147 | (0%) |
| Down’s syndrome | 4 | (0%) | 16 | (0%) | 7 | (0%) |
| Family history of cardiovascular disease | 239 | (0%) | 1,636 | (1%) | 1,778 | (1%) |
| **Medication history [n (%)]** |  |  |  |  |  |  |
| Antihypertensive drugs | 25,986 | (53%) | 102,429 | (73%) | 87,215 | (73%) |
| Antidiabetic drugs | 8,923 | (18%) | 36,480 | (26%) | 31,860 | (27%) |
| Antiplatelet drugs | 16,882 | (35%) | 61,991 | (44%) | 51,168 | (43%) |
| Non-steroidal anti-inflammatory drugs | 14,018 | (29%) | 58,562 | (42%) | 57,593 | (48%) |
| Corticosteroids | 15,391 | (32%) | 68,034 | (48%) | 58,831 | (49%) |
| Proton-pump inhibitors | 8,666 | (18%) | 33,265 | (24%) | 26,564 | (22%) |
| H2-receptor antagonists | 16,454 | (34%) | 76,727 | (55%) | 68,887 | (58%) |
| Anticoagulants | 2,994 | (6%) | 7,329 | (5%) | 7,001 | (6%) |
| Nicotine replacement therapy | 386 | (1%) | 966 | (1%) | 1,920 | (2%) |
| Antiarrhythmic drugs | 1,269 | (3%) | 3,194 | (2%) | 2,462 | (2%) |
| Antithyroid drugs | 325 | (1%) | 1,469 | (1%) | 1,290 | (1%) |
| Oestrogen | 358 | (1%) | 652 | (0%) | 534 | (0%) |
| Psychotropic drugs | 7,013 | (14%) | 24,838 | (18%) | 23,931 | (20%) |
| Cardiac glycosides | 1,498 | (3%) | 5,397 | (4%) | 3,561 | (3%) |
| Nitrates | 10,250 | (21%) | 40,312 | (29%) | 29,230 | (24%) |
| Thyroid hormones | 1,208 | (2%) | 3,810 | (3%) | 3,238 | (3%) |
| Testosterone | 226 | (0%) | 731 | (1%) | 922 | (1%) |
| Fibrates | 1,997 | (4%) | 8,252 | (6%) | 6,126 | (5%) |
| Niacin | 72 | (0%) | 94 | (0%) | 67 | (0%) |
| PCSK9 inhibitors | 3 | (0%) | 3 | (0%) | 9 | (0%) |
| Cholesterol absorption inhibitors | 179 | (0%) | 340 | (0%) | 379 | (0%) |
| Vytorin | 3 | (0%) | 1 | (0%) | 0 | (0%) |
| Bile acid sequestrants | 118 | (0%) | 156 | (0%) | 78 | (0%) |
| Omega-3 fatty acids | 28 | (0%) | 11 | (0%) | 3 | (0%) |
| Other non-statin lipid-modifying drugs | 1 | (0%) | 0 | (0%) | 5 | (0%) |
| **General (**before incident cardiovascular events**) [median (interquartile range)]** | | | | | | |
| Outpatient visits per year | 3.0 | (0.0-4.6) | 5.3 | (2.3-7.2) | 4.9 | (2.2-6.5) |
| Inpatient visits per year | 0.8 | (0.8-0.8) | 0.9 | (0.7-1.0) | 0.9 | (0.7-0.9)* |
| Count of medications | 5.0 | (0.0-8.0) | 7.0 | (5.0-10.0) | 7.0 | (5.0-10.0) |
| PCSK9 = Proprotein convertase subtilisin/kexin type 9. H2 = histamine type 2.  *Risk variables in the Kowloon and New Territories cohorts with no significant difference in distribution (p value≥0.05) from the Hong Kong Island (Hong Kong West Cluster) under Chi-square test (categorical risk variables) or in T-test (numerical risk variables). All other risk variables were significant (p value<0.05). | | | | | | |

**Supplementary Information 1. Details of data source**

The Hong Kong Island (Hong Kong West Cluster) cohort was identified by the Hospital Authority, which included all patients of age 18 or above at the time when they received their lipid test at the hospitals located in Hong Kong West Cluster between 1 January 2004 and 31 December 2019. P-CARDIAC is derived from the Hong Kong Island (Hong Kong West Cluster) cohort.

For the Kowloon and New Territories cohorts, a 2 million patient cohort was retrieved from the Hospital Authority database. Any patients aged 35-year or above at the time when they had their blood pressure recorded in the Hospital Authority between 1 January 2005 and 31 December 2019. External validation was completed using the Kowloon and Kew Territories cohorts to ensure no overlap with the model derived cohort.

**Supplementary Information 2. Gradient boosting Cox proportional hazards modeling using XGBoost**

The Cox model is expressed by the hazard function denoted by h(t). Briefly, the hazard function can be interpreted as the risk of dying (or other event of interest happening) at time t.

ln(h(t)) = ln(h0(t)) + <w,x>

Where:

x is a vector in Rd representing the features.

w is a vector consisting of d coefficients, each corresponding to a feature.

⟨⋅,⋅⟩ is the usual dot product in Rd.

ln(⋅) is the natural logarithm.

the term h0(t) is the baseline hazard.

XGBoost revises the model as follows to make Cox work with gradient boosting:

ln(h(t)) = ln(h0(t)) + T(x)

where T(x) represents the output from a decision tree ensemble, given input x. The goal for XGBoost is to maximize the (log) likelihood by fitting a good tree ensemble T(x).

**Supplementary Information 3. Design of the hinge loss-like function**

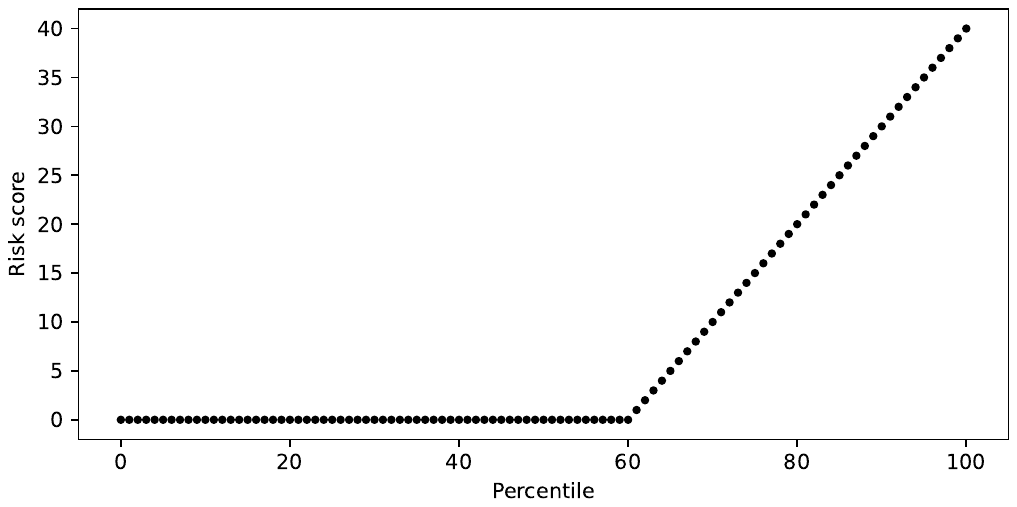

f(p) = max(0, p-t)

t is the threshold with value of 60

p is the discrete percentile of the hazard ratio for all involved patients.

**Supplementary Information 4. Screenshots and clinical example of the P-CARDIAC website interface**

1. Fill in mandatory fields

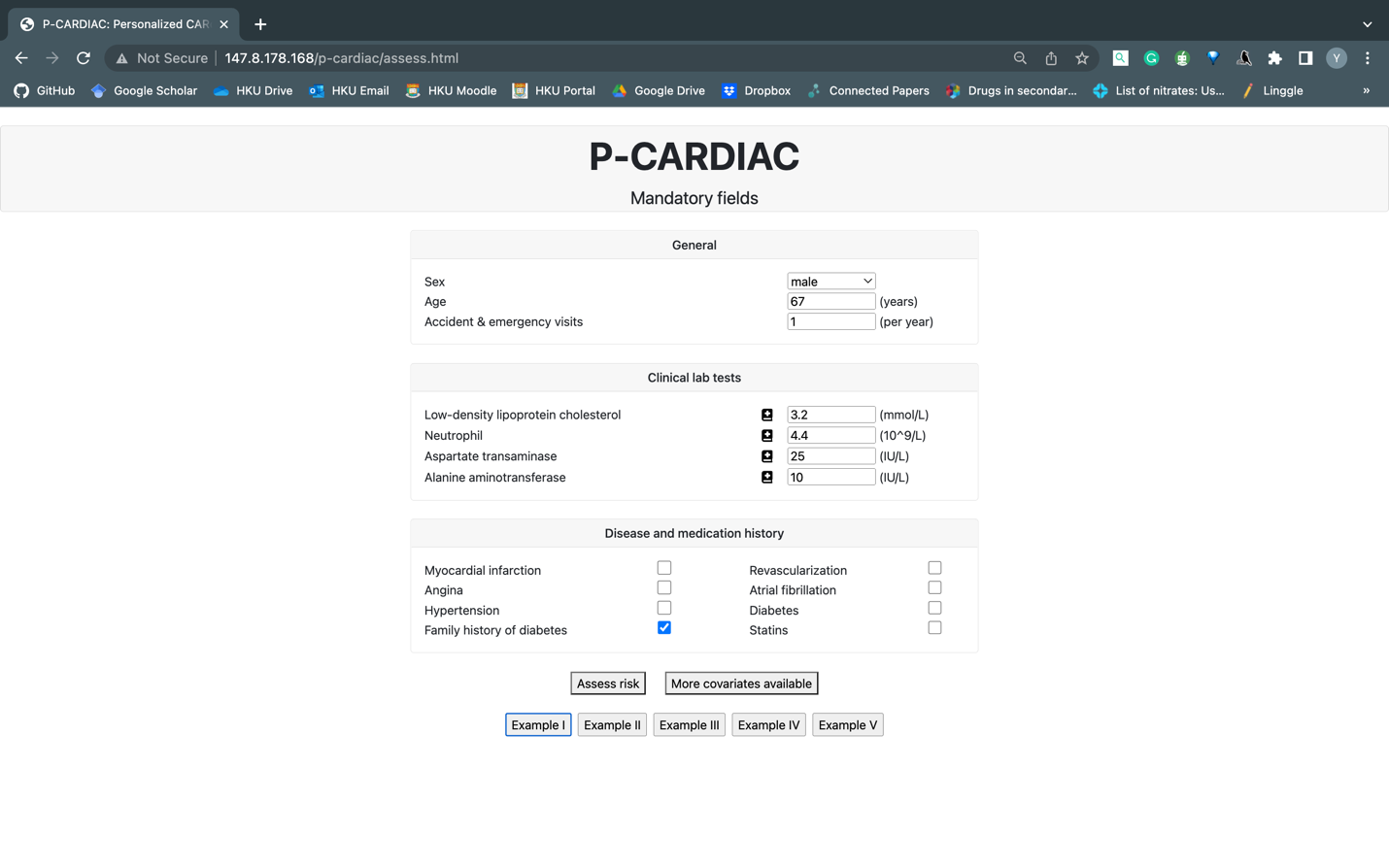

2. Assess risk based on only mandatory risk variables (predicted 10-year CVD risk of 91.0%)

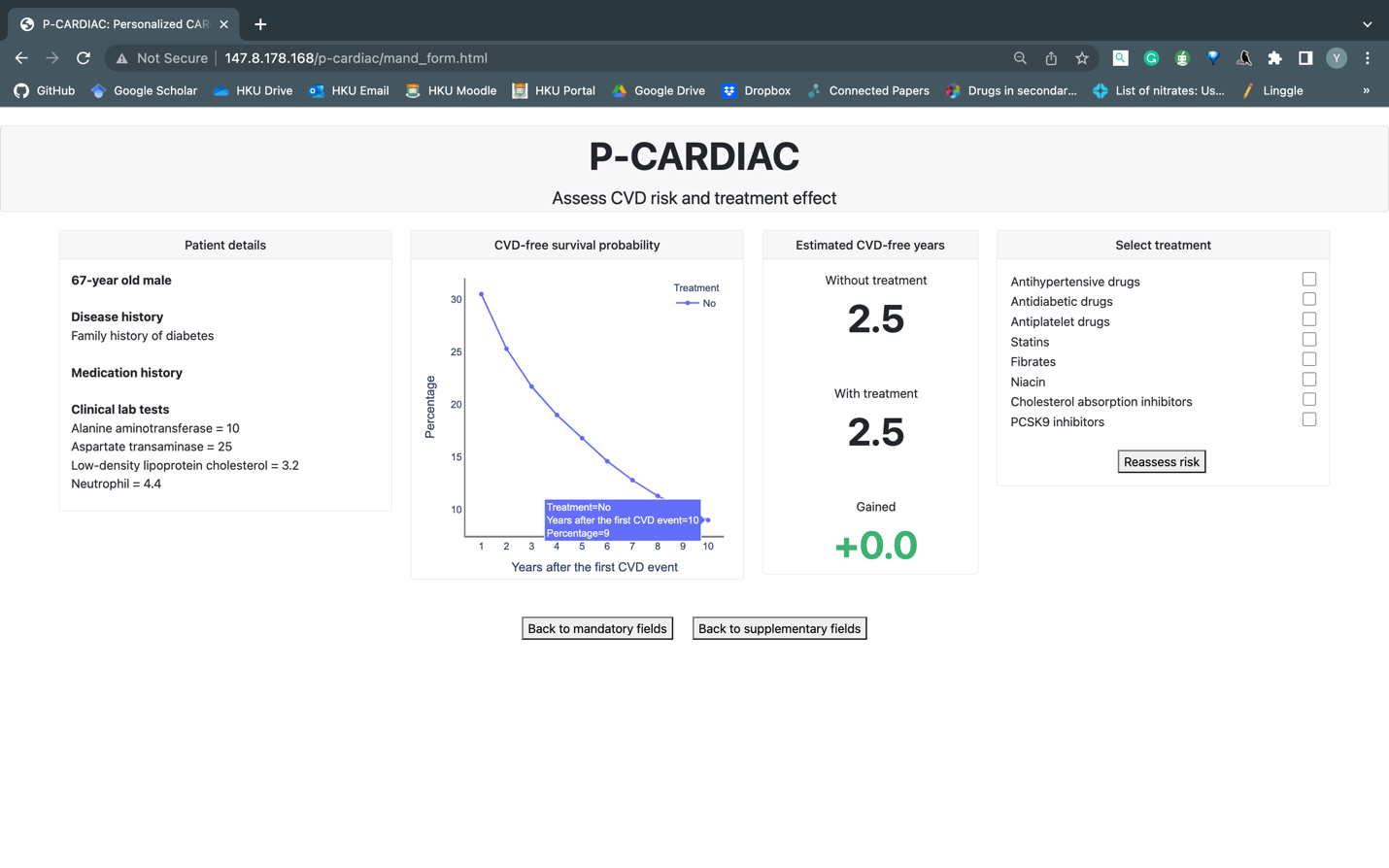

3. Fill in supplementary fields

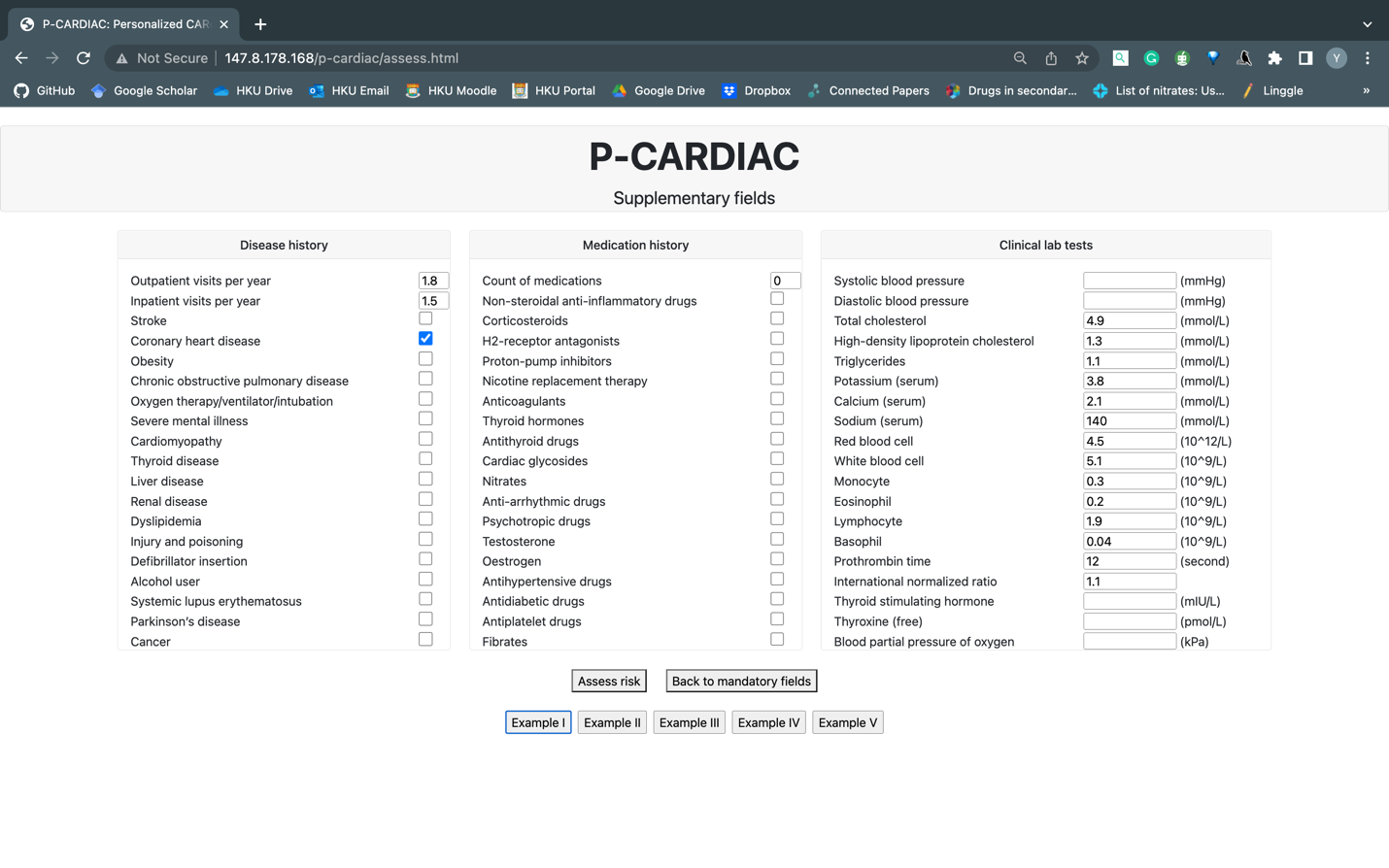

4. Assess risk based on both mandatory and supplementary risk variables (predicted 10-year CVD risk of 93.9%)

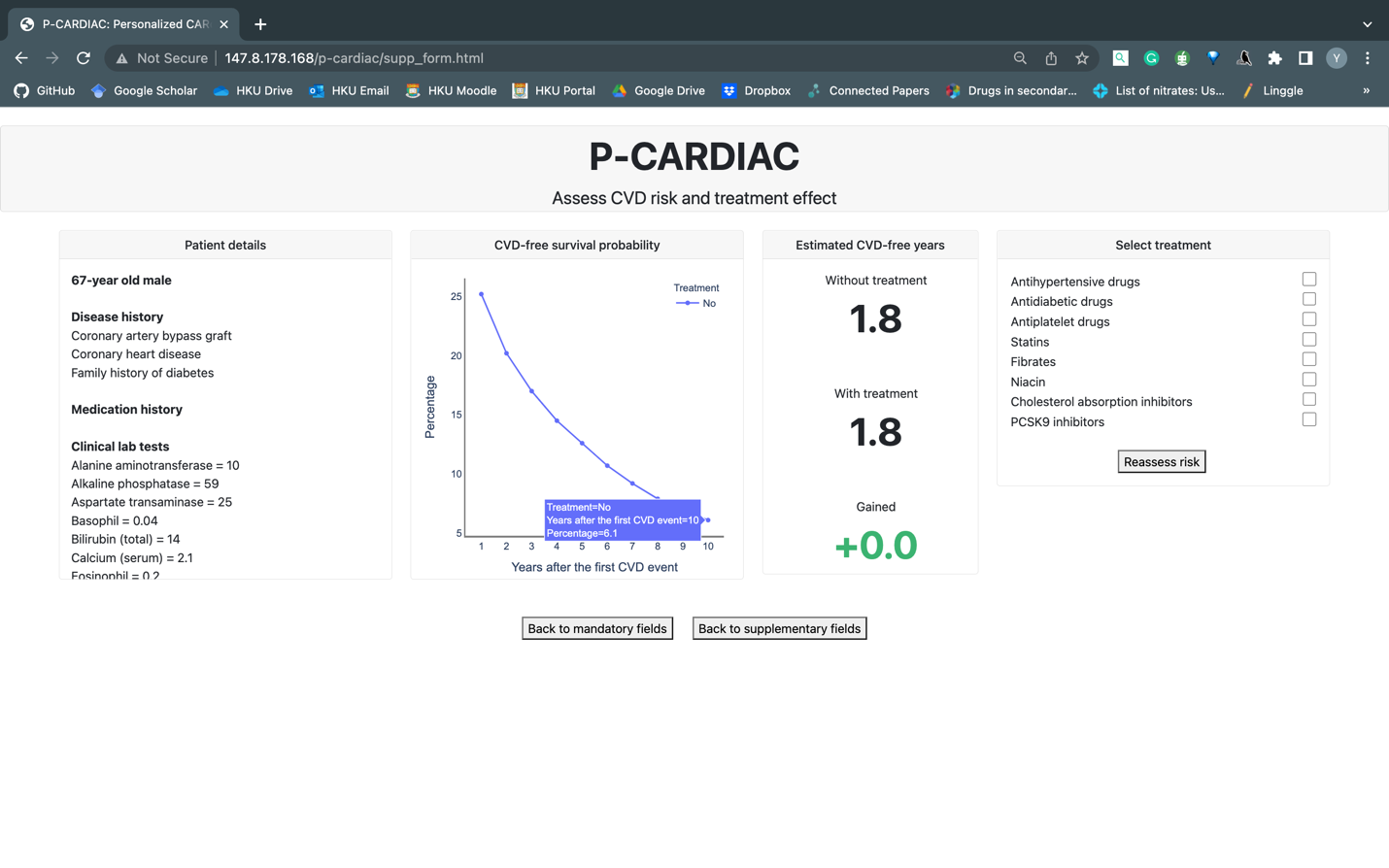

5. Add statins to treatment (predicted 10-year CVD risk of 88.2%)

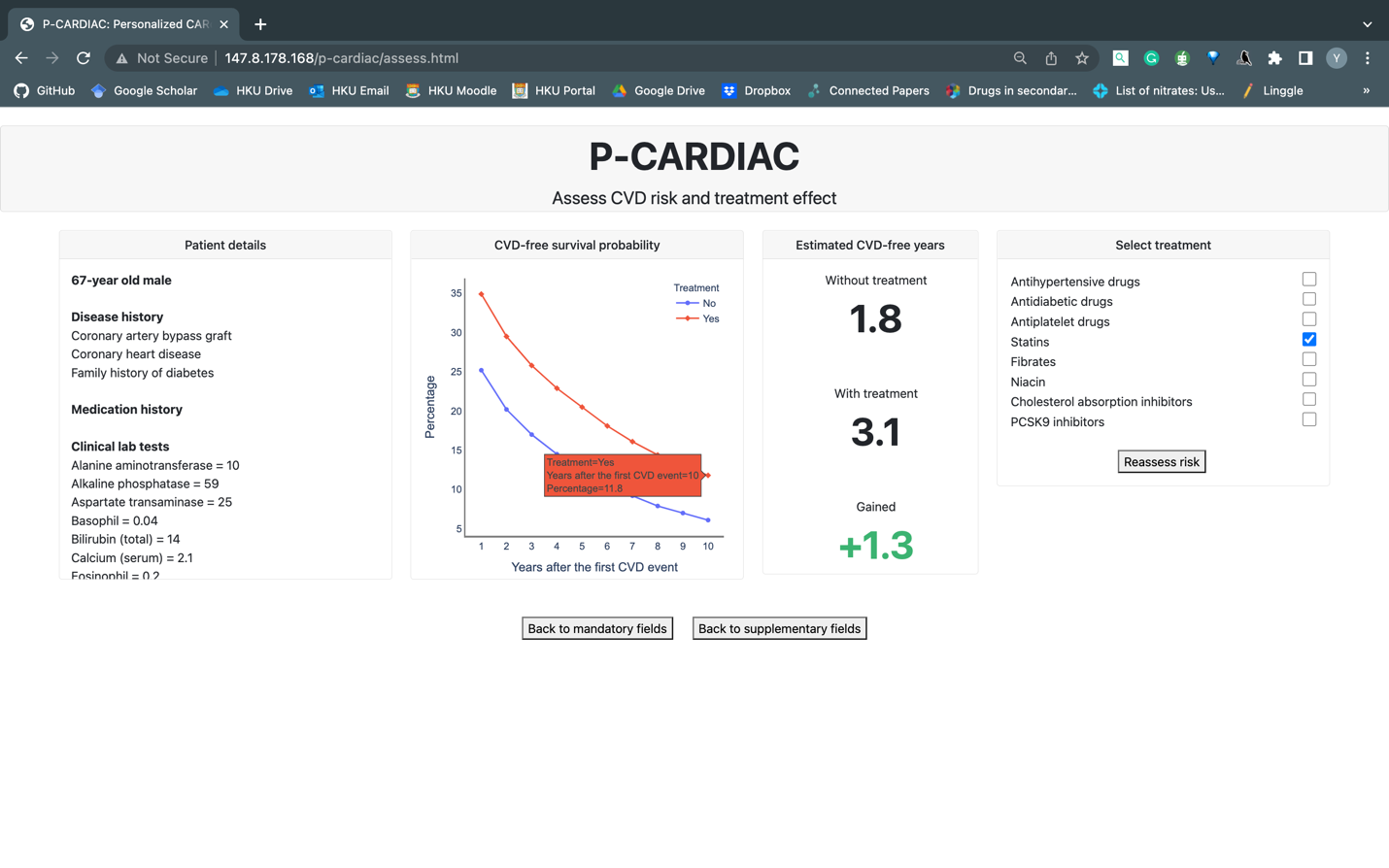

6. Add PCSK9 inhibitors to treatment (predicted 10-year CVD risk of 41.4%)

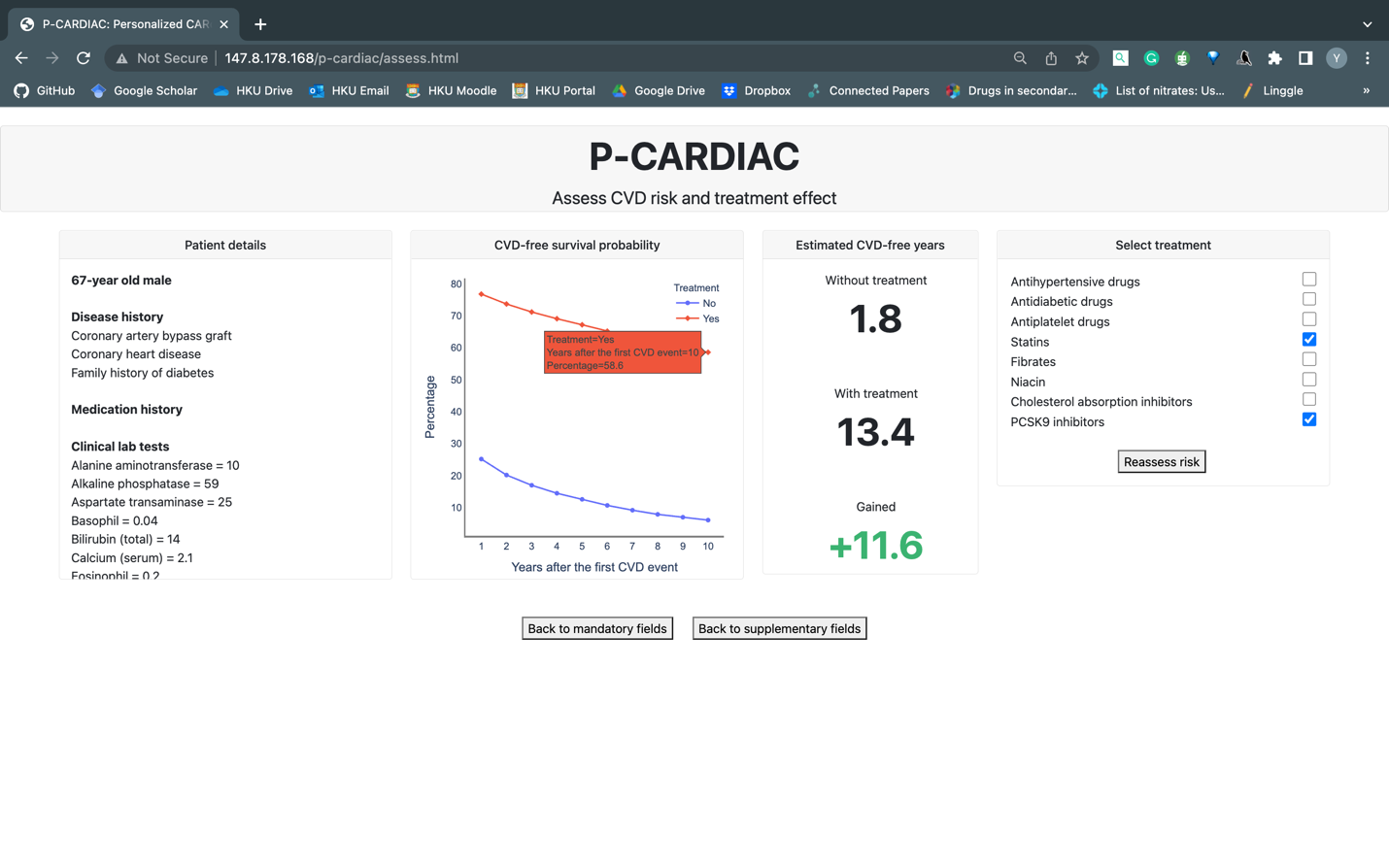
